# Calibrated Prediction Intervals for Polygenic Scores: Updated Comparisons, Contextual Calibration, and Data Normalization

**DOI:** 10.64898/2026.05.15.26353336

**Authors:** Chang Xu, Siyu Hou, Xiang Zhou

**Author notes:** authors contributed equally (Chang Xu, Siyu Hou).

## Abstract

Calibrated prediction intervals for polygenic scores (PGS) are essential for communicating individual-level uncertainty in genomic medicine. We present updated comparisons of two methods for constructing such intervals: CalPred, a parametric approach, and PredInterval, a non-parametric approach. Our results show that both methods can achieve calibrated coverage, although CalPred additionally requires a sufficiently large calibration set. The two methods also exhibit complementary trade-offs with respect to dataset size and risk identification. We further show that contextual calibration, as introduced in Hou et al. and followed in Shi et al., is most naturally achieved through appropriate phenotype normalization and data preprocessing. Apparent miscalibration can arise from inadequate normalization or from providing contextual information to some methods but not others. In UK Biobank, standard GWAS phenotype normalization procedures are sufficient to achieve contextual calibration for traits analyzed. In the extreme simulations of Hou et al. and Shi et al., supplying contextual covariates to PredInterval restores contextual calibration without normalization, and appropriate normalization can achieve contextual calibration without supplying covariates, while also substantially improving upstream tasks including association power and PGS accuracy. Together, these results underscore the central role of phenotype normalization and data preprocessing in GWAS analyses, including reliable uncertainty quantification for PGS.

## Main

Polygenic scores (PGS) summarize an individual’s genetic predisposition to complex traits and diseases and are increasingly used in research and clinical settings[1]. However, point estimates alone do not communicate the uncertainty in individual-level predictions. Prediction intervals are therefore essential for responsible use, enabling calibrated risk communication and helping distinguish truly high-risk individuals from those near the population average. Calibration is central: if nominal coverage is not achieved in practice, intervals may overstate certainty, which risks inappropriate decisions, or understate it, which reduces clinical utility.

### Marginal calibration in practice: calibration-set size and real-data performance

Two recently proposed methods address this need: CalPred[2], a parametric approach operating on a scaled PGS based on a heteroskedastic regression model, and PredInterval[3], a non-parametric approach operating on the original PGS based on prediction residuals. Following kind clarifications from the team of Shi et al.[4] about CalPred’s intended fitting procedure, which resolves ambiguity in the public software documentation (details in Supplementary Note), we reanalyzed our previous simulations and confirm that CalPred attains coverage centered at the target level in simulations (Fig. 1a) and shows improved performance for identifying high-risk individuals, approaching PredInterval (Fig. 1b).

**Figure 1.**
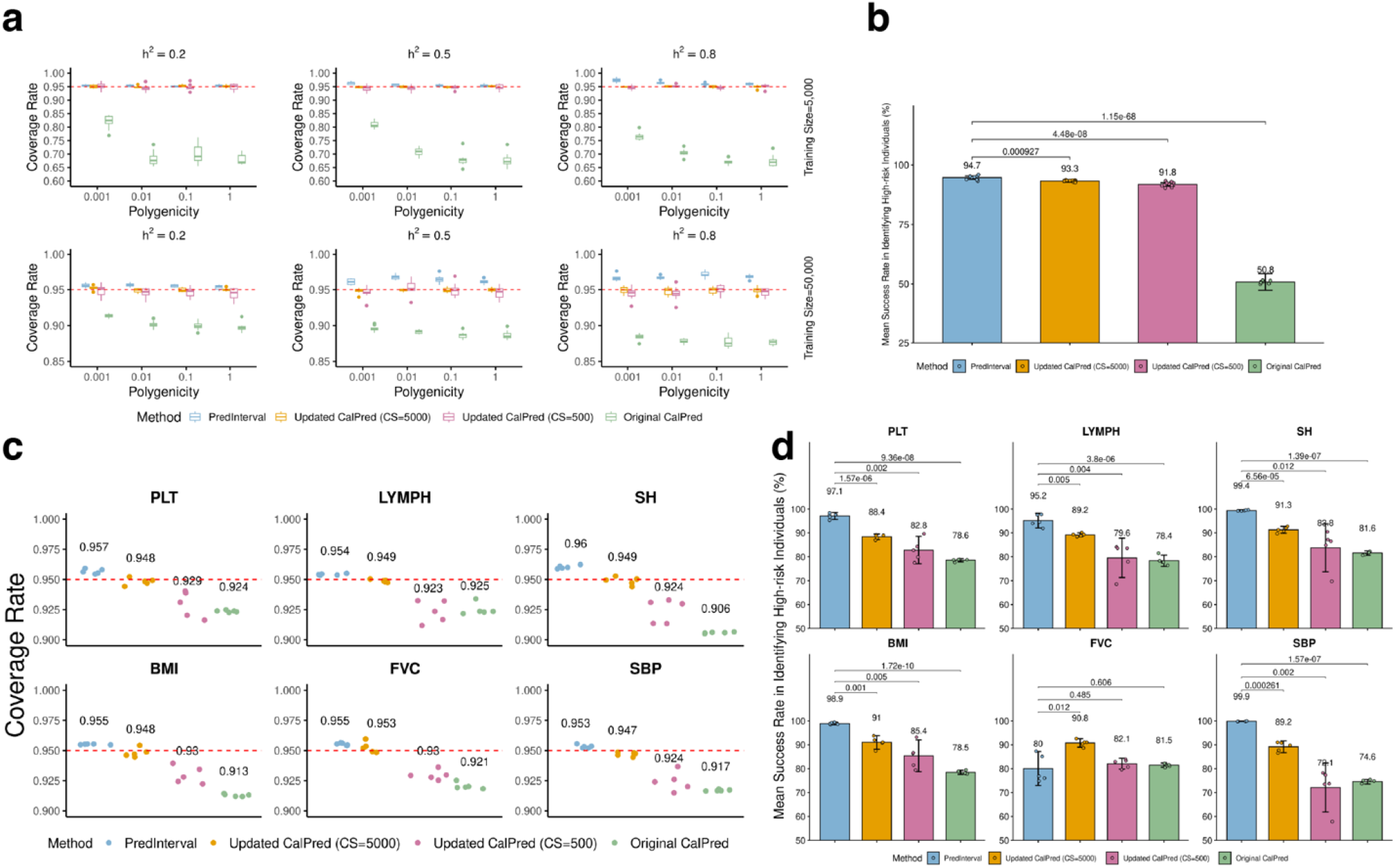
Updated marginal calibration comparison results in simulations and real data. Compared methods include the originally fitted CalPred, updated CalPred with calibration size of 500 (cs = 500), updated CalPred with calibration size of 5,000 (cs = 5,000), and PredInterval. Results are shown for (a) prediction interval coverage rate and (b) individual risk stratification in simulations, as well as (c) prediction interval coverage rate and (d) mean success rate in individual risk stratification across six traits in the UK Biobank applications in real data.

In real data, however, correcting the fitting procedure alone does not fully resolve CalPred’s downward-biased coverage (Fig. 1c), consistent with the close agreement between results in Xu et al. and Hou et al. By carefully examining the CalPred implementation used in Shi et al. to pinpoint the source of the discrepancy, we found that Shi et al. used a substantially larger calibration set, which is over 100x larger than the 500-sample recommended in Hou et al. Motivated by this difference, we also found that increasing the calibration set size improves coverage of CalPred: its coverage is centered near the nominal level when the calibration set is roughly ten times larger than the recommendation. However, even with updated usage and a ten-fold larger calibration set, CalPred’s high-risk identification performance remains lower for most traits (Fig. 1d), except for forced vital capacity (FVC), where sex-specific phenotypic variance is not addressed by simple normalization, as we discuss below.

### Contextual calibration in real data: standard data normalization is sufficient

Contextual calibration, introduced in Hou et al. and followed in Shi et al., refers to achieving nominal prediction-interval coverage not only marginally but also within strata defined by covariates such as sex and age. Contextual miscalibration can arise when covariate effects induce differences in the phenotype’s variance in addition to the mean across strata, and when these effects are not adequately removed during data normalization and preprocessing. This can occur in several cases: (i) incomplete adjustment of non-linear mean effects (e.g., adjusting for age but not age^2^), which can leave structured residual variation across strata; (ii) genuine variance heterogeneity across covariate strata (e.g., differing phenotypic variance between males and females) but failing to adjust them during data normalization; and (iii) differences in covariate effects between training and test data (e.g., because they come from distinct studies or populations), in which case the held-out data used by PredInterval or the calibration samples used by CalPred, if not drawn from the test distribution, may no longer be representative of the test data, inducing miscalibration. In all cases, appropriate data normalization and preprocessing is the most direct and effective remedy (details in Methods).

Standard normalization, regressing out covariates and applying inverse-quantile normalization to the residual phenotype, is routine in GWAS. When this preprocessing is properly applied, we find that PredInterval, even without explicitly modeling covariates, achieves contextual calibration for the majority of analyzed traits in UK Biobank across both sex and age strata (Fig. 2a–b). The only exception is forced vital capacity (FVC), which shows minor sex-stratified deviation (coverage 0.93 in males and 0.97 in females vs. target 0.95). This pattern is consistent with sex-specific variance heterogeneity (variance 1.31 in males vs. 0.74 in females) and can be addressed by applying quantile normalization separately within each sex (Fig. 2c), aligning with common practice for this trait (within-sex normalization or sex-stratified analyses)[5].

**Figure 2.**
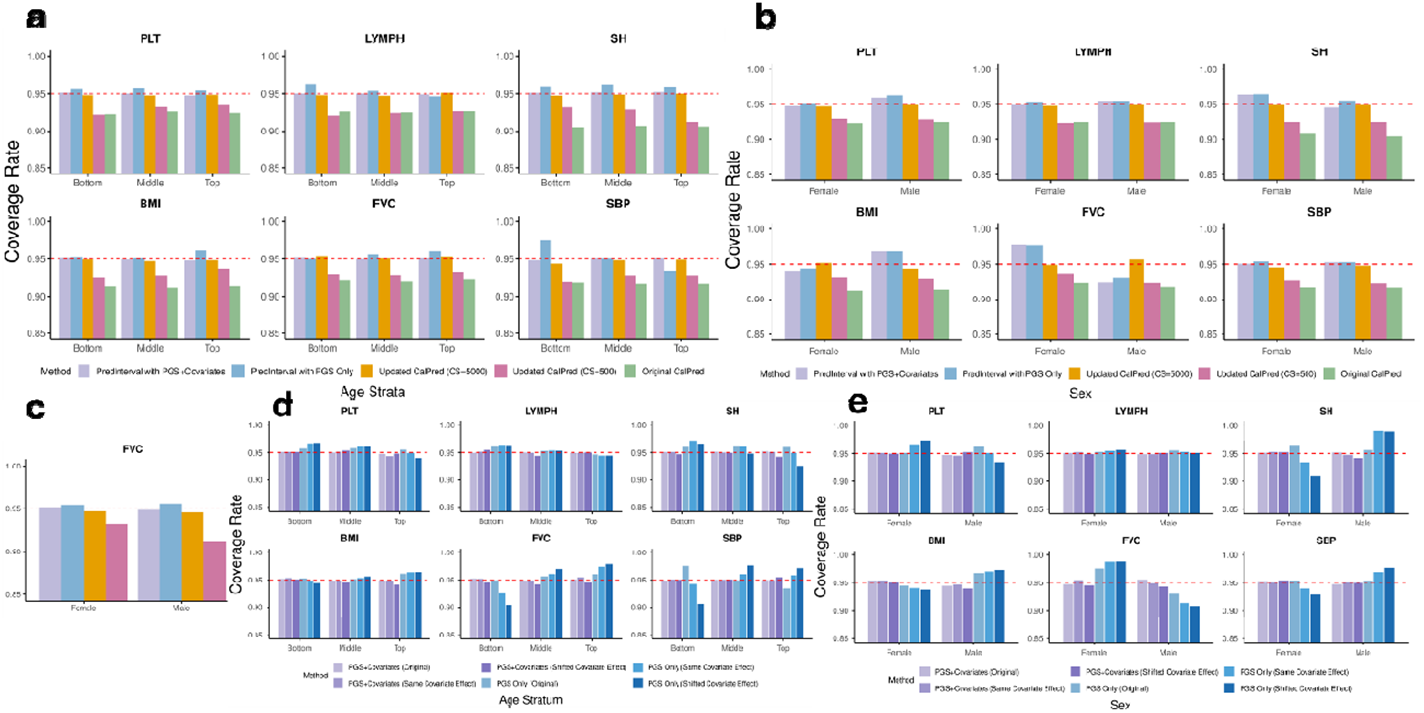
Examining contextual calibration in UK Biobank. Compared methods include the originally fitted CalPred, updated CalPred with calibration size of 500 (cs = 500), updated CalPred with calibration size of 5,000 (cs = 5,000), PredInterval with PGS only (the original version), and PredInterval with PGS + Covariates (the extended version that explicitly models covariate effects). Results are shown for contextual calibration across sex and age strata for six traits in the UK Biobank. (a)–(b): Results using the originally reported phenotypes, where covariate effects were controlled during standard phenotype normalization in the original analysis. PredInterval with PGS only already achieves contextual calibration for the majority of analyzed traits under this normalization. (c): Results for FVC using sex-stratified normalization, where quantile normalization is performed separately within each sex to address residual variance heterogeneity between males and females. (d)-(e): Results for two versions of PredInterval (PGS Only, and PGS + Covariates), using either the original phenotype, or two sets of reconstructed phenotypes, where the original standardized effects of sex, age, and age^2^ are added back to the normalized phenotype either consistently across all samples (Same Covariate Effect), creating a setting in which covariate effects are not properly adjusted, or with a larger magnitude in the test data than in the training data (Shifted Covariate Effect), thereby inducing a mismatch in covariate effects between training and test sets.

To illustrate the impact of inadequate data preprocessing, we reconstructed an “unnormalized” phenotype by adding sex, age, and age^2^ effects back to the normalized phenotype, creating a setting where covariate effects remain in the data. Contextual miscalibration becomes more pronounced, with additional traits showing miscoverage across sex and age strata. For example, standing height (SH) becomes miscalibrated with respect to both sex and age, in addition to the sex-related miscalibration for FVC (Fig. 2d–e). We also considered a more extreme distribution-shift setting in which sex, age, and age^2^ effects in the test set are twice those in training, mimicking scenarios where training and test samples differ in covariate distributions or measurement protocols – for example, when they come from distinct studies or populations. Miscalibration increases substantially and affects a broader set of traits (including PLT, BMI, FVC, and SH) across both sex and age strata (Fig. 2d–e), at a magnitude consistent with what was observed in Shi et al.’s cross-study UK Biobank/All of Us analysis.

Together, these results indicate that contextual miscalibration is primarily a preprocessing problem: contextual calibration generally requires removing (or correctly modeling) covariate effects before constructing intervals. For rarer settings where proper normalization is not feasible or has not been performed, we previously developed an extension of PredInterval that explicitly incorporates covariate effects on both the mean and the variance. A detailed software manual was provided on GitHub along with brief modeling descriptions available on GitHub and in the code annotations; here we provide full technical details in the Methods. The extension preserves PredInterval’s nonparametric framework and uses a two-stage procedure to model covariate effects on both the mean and residual scale central to PredInterval’s construction. This covariate-aware PredInterval achieves contextual calibration in settings where normalization was not performed (Fig. 2a–e), including under train–test shifts in covariate effects, in which a portion of the test data should serve as the held-out set (for PredInterval) or the calibration set (for CalPred).

### Contextual miscalibration in simulations arises from unnormalized phenotypes and omitted context

Because we do not observe contextual miscalibration issues in real data, we ask why Hou et al. and Shi et al. observed contextual miscalibration for other methods in the simulations. To understand the findings in these two studies, it is helpful to examine how their simulations were generated and how the subsequent analyses were conducted. Specifically, both studies generate outcomes under extreme contextual heterogeneity using a data-generating strategy fully based on the parametric CalPred model. For example, the most extreme setting in Shi et al. simulates heteroskedastic noise whose variance is an exponential function of a normally distributed contextual variable, so that the residual variance in the top decile of the context (=6.416) is 34.56-fold higher than in the bottom decile (=0.186). This is further combined with homogeneous genetic effects completely independent of the context and are unrealistically large: it has a polygenic architecture in which only 82K SNPs explain 50% of phenotypic variance (*h*^2^ = 0.5), and the resulting genomic-control factor in 60K samples has a median of 6.835 (range: 6.547–7.080) across replicates. The genetic component is then rescaled to have mean zero and standard deviation one before being added to the residual to form the simulated trait, yielding extreme context-dependent heritability: *h*^2^ = 0.13 in the top decile of the context and *h*^2^ = 0.84 in the bottom decile. Importantly, both studies analyze the simulated data without appropriate data normalization, while providing the contextual variable to CalPred but not to competing methods.

In our re-analysis of the most extreme setting in Shi et al., we similarly find that omitting normalization induces contextual miscalibration for PredInterval when contextual information is not supplied (Fig. 3a and 3c). However, different from Shi et al., we find that supplying the contextual variable to PredInterval restores contextual calibration even without normalization (Fig. 3a and 3c), consistent with the robustness expected of a nonparametric approach and its effectiveness even when model assumptions do not match the data-generating process. More importantly, we show that standard normalization for heterogeneous noise (e.g., two-stage linear regression; Methods) is sufficient to achieve contextual calibration even without providing context to PredInterval when the genetic effects are large but not completely unrealistic (*h*^2^ = 0.05 by 82K SNPs; median genomic-control factor was 1.540, range 1.474–1.654; Fig. 3a). Even under the unrealistically large genetic effect setting (*h*^2^ = 0.5 by 82K SNPs), normalization applied to estimated residuals still yields contextual calibration (Fig. 3c).

**Figure 3.**
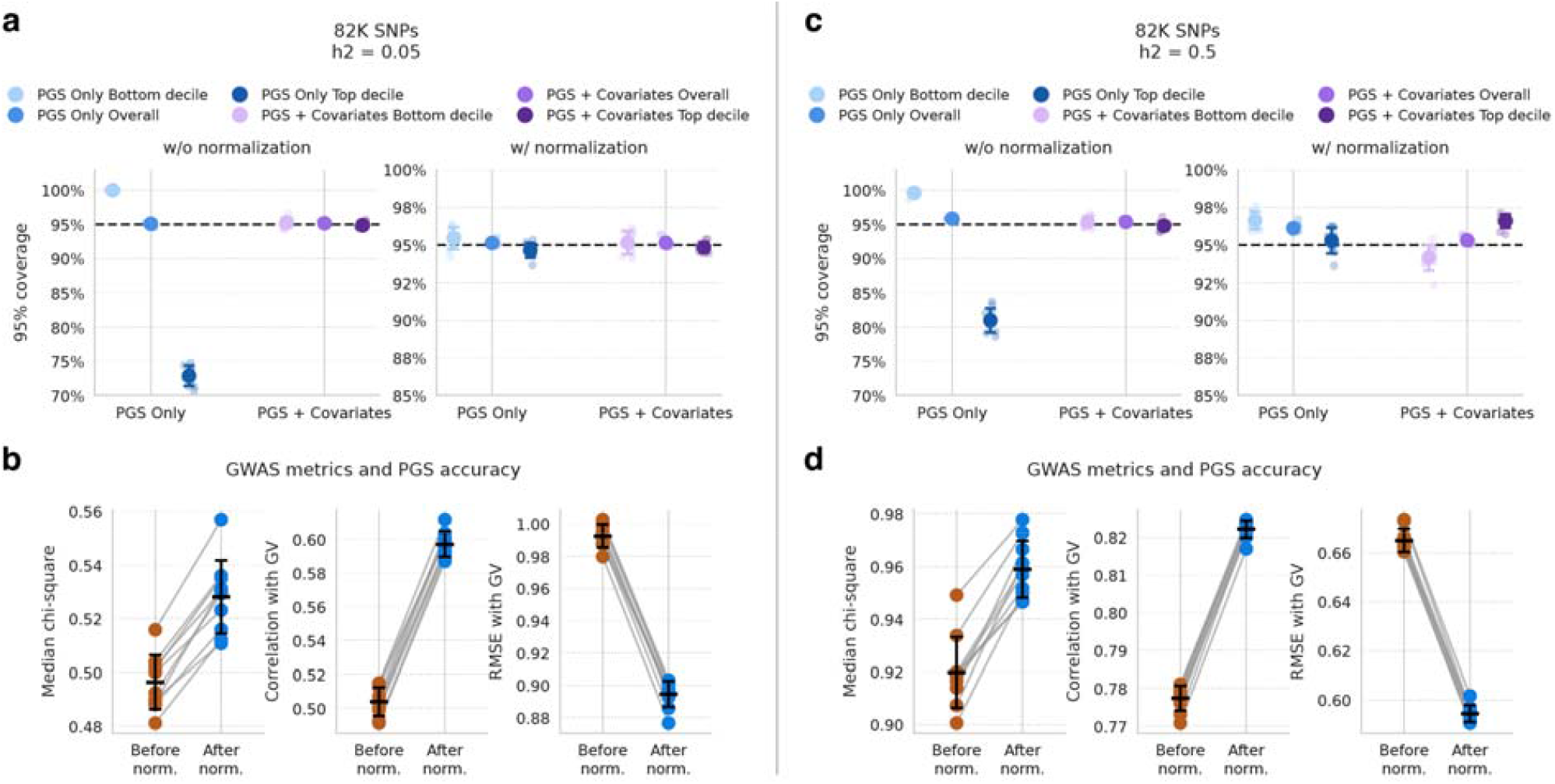
Proper phenotype normalization is key to achieving contextual calibration, improving upstream association performance, and increasing PGS accuracy in the most extreme simulation setting of Shi et al. Results are shown for two simulation settings, *h*^2^ = 0.05 (a, b) and *h*^2^ = 0.5 (c, d). In each setting, we compared analyses using the raw phenotype versus the normalized phenotype, including prediction-interval calibration (a, c), GWAS signal as summarized by the median *χ*^2^ statistic (b, d), and PGS accuracy measured by correlation with (or RMSE against) the true genetic value (b, d), all evaluated in the test set. For prediction-interval calibration, we applied two versions of PredInterval: PredInterval using PGS only, and PredInterval using PGS plus covariates.

Because normalization reduces heteroskedastic noise, it is especially crucial for valid upstream analytic tasks, including association testing and PGS construction, since standard models for both tasks typically assume homoskedastic errors. Consistent with this, normalization substantially improves upstream association results and prediction in simulations. For the *h*^2^ = 0.05 setting, SNP association signal increases: the median chi-square statistics across causal SNPs rises from 0.496 (range 0.481-0.516 across replicates, evaluated in the 10K sample test set) with the unnormalized phenotype to 0.528 (range 0.511-0.557) after normalization (Fig. 3b). PGS accuracy also improves: the median correlation with the true genetic value increases from 0.503 (range 0.492-0.511, evaluated in the test set) to 0.597 (range 0.587-0.611), and RMSE decreases from 0.993 (range 0.983-1.002) to 0.896 (range 0.877-0.903) (Fig. 3b). Even under the unrealistically large genetic effect setting (*h*^2^ = 0.5 by 82K SNPs), applying normalization to estimated residuals substantially improves both association signal detection and PGS accuracy: the median chi-square statistics increases from 0.920 (range 0.901-0.949) to 0.959 (range 0.946-0.978) (Fig. 3d), the correlation with the true genetic value improves from 0.778 (range 0.771-0.781) to 0.823 (range 0.817-0.825), and RMSE decreases from 0.663 (range 0.660-0.673) to 0.595 (range 0.591-0.602) (Fig. 3d).

## Discussion

We present updated comparisons of CalPred and PredInterval and show that, when applied as intended, both methods yield well-calibrated prediction intervals, including contextual calibration. For CalPred, achieving nominal coverage additionally requires a sufficiently large calibration set. More broadly, our results highlight that phenotype normalization and preprocessing are central to contextual calibration and can simultaneously and substantially improve upstream analyses, including association power and PGS accuracy.

Taken together with Xu et al., these results emphasize practical trade-offs between the two approaches. CalPred can underestimate coverage when the calibration set is small, but approaches nominal coverage with sufficiently large calibration data; in our UK Biobank application, roughly ten times the default recommendation was sufficient, though the required size may vary by dataset. PredInterval, by contrast, does not require a minimum calibration-set size because coverage is guaranteed by construction under exchangeability; however, it can be conservative when a large portion of training data is withheld to compute residual-based nonconformity scores, and converges toward nominal coverage as the withheld subset shrinks. These differences also affect high-risk identification: PredInterval’s conservatism yields wider intervals and greater sensitivity for identifying high-risk individuals, potentially at the cost of more false positives, whereas CalPred, when ample calibration data are available, tends to produce tighter intervals with fewer false positives. Part of this contrast reflects that the methods target different predictors: PredInterval constructs intervals around the user-supplied (original) PGS, while CalPred effectively rescales (and often shrinks) the PGS by regressing the phenotype on it, producing intervals around that scaled prediction. Accordingly, PredInterval can be used either with the original PGS or with a rescaled PGS supplied by the user when comparability across settings is desired.

Our analyses also clarify that contextual miscalibration is typically not an inherent limitation of the interval-construction methods. Instead, it most often reflects differences in preprocessing and/or differences in whether contextual variables are provided when contextual calibration is evaluated. In UK Biobank, standard GWAS-style preprocessing achieves contextual calibration for the traits analyzed, with a minor deviation for one trait consistent with sex-specific variance heterogeneity that is corrected by stratified normalization. In the extreme simulation settings considered by Hou et al. and Shi et al., contextual calibration is recovered either by providing contextual covariates to methods designed to use them or by applying appropriate normalization; importantly, normalization also improves upstream association testing and PGS construction. When normalization is not feasible or has not been performed, explicit modeling of covariate effects on both the mean and variance scales becomes necessary, and our covariate-aware extension of PredInterval provides a principled nonparametric option in these settings.

Finally, this experience highlights broader lessons for computational genomics. Software documentation is the primary interface between a method and its users, and clear documentation, together with careful adherence to the software manual, is often essential to reproduce intended behavior. In retrospect, our use of CalPred in Xu et al. and the use of the covariate-aware version of PredInterval in Shi et al. each followed the available software instructions, yet differed from the developers’ intended usage in ways that affected marginal and contextual calibration, respectively. Resolving these discrepancies required additional communication between groups – an effort that could often be avoided with clearer software documentation and more explicit guidance on recommended workflows. We view this process as a constructive example of how transparency and open exchange strengthen methodological science.

## Methods

### Updated CalPred fitting for marginal calibration in simulations and real data applications

All simulations and real data applications for marginal calibration follow the setup described in Xu et al. Per the kind recommendation from the team of Shi et al., we updated the CalPred fitting across all simulations and real data applications for quantitative traits. We followed the CalPred paper’s recommendation and set the calibration set size to 500, and additionally explored a setting ten times larger at 5,000 following careful examination of Shi et al..

To facilitate the use of existing fitted models, calibration sets were carved out from the original test set. In simulations, we randomly sampled individuals from the original test set of 10,000 to form the calibration set of the specified size, and treated the remaining individuals as the new test set for CalPred. In real data applications, for each fold of the five-fold cross-validation, we similarly split the calibration set from the original test set (∼72,000 individuals) and treated the remaining individuals as the new test set. In both settings, PGS values in the calibration and test sets were constructed using SNP weights estimated by DBSLMM in the original training set, CalPred was fitted on the calibration set, and prediction intervals were generated for the test set.

### Sources of contextual miscalibration and corresponding phenotype normalization and data preprocessing remedies

Contextual calibration refers to achieving nominal prediction-interval coverage not only marginally, but also within strata defined by covariates (e.g., sex, age groups). Contextual miscalibration can arise when covariates induce differences across strata in the phenotype’s distribution, most commonly in the mean, but sometimes also in the variance, and these effects are not adequately addressed during phenotype preprocessing. Below we distinguish three practically important scenarios and summarize the standard statistical remedies.

#### Case I: Nonlinear mean effects that are not fully adjusted

A common source of apparent “variance differences” across context is incomplete adjustment of nonlinear mean structure. For example, adjusting for age linearly while omitting age^2^, or other nonlinear terms and interactions, can leave systematic residual patterns that differ across covariate strata and later appear as contextual miscalibration. In our experience, standard GWAS-style mean adjustment, through regressing the phenotype on covariates with a sufficiently flexible specification (e.g., including age^2^) and applying inverse-quantile normalization to the residuals, is often sufficient in this setting.

#### Case II: Genuine variance heterogeneity across strata (heteroskedasticity)

For some traits, covariates affect not only the mean but also the variance. For example, forced vital capacity (FVC) may be intrinsically more variable in one sex than the other. When such heteroskedasticity is present, mean-only normalization can leave residual variance differences across strata, which can sometimes translate into observable contextual miscalibration of prediction intervals. In this setting, additional preprocessing beyond mean adjustment may be required.

Many standard data normalization approaches can address heteroskedasticity. One simple option is a two-stage regression procedure. In the first stage, we fit a mean model *m* (·) and compute residuals 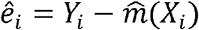. In the second stage, we model residual scale as a function of covariates, for example, by regressing 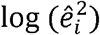 on *X*_*i*_, yielding an estimated conditional scale 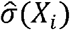. We then define a variance-stabilized phenotype 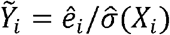 optionally followed by inverse normalization.

An alternative, especially natural when the primary driver of heteroskedasticity is a discrete covariate (e.g., sex), is stratum-wise quantile normalization: we apply inverse normalization separately within each stratum so that the within-stratum phenotype distribution is comparable across strata. Both strategies aim to remove variance differences across contexts so that downstream uncertainty quantification targets a stable, context-invariant residual scale.

#### Case III: Train-test differences in covariate effects (distribution shift)

Even with careful normalization, contextual miscalibration can occur under train-test shift when covariate effects or other aspects differ between training and test samples (e.g., due to different studies, recruitment mechanisms, or populations). A common example is training a model in UK Biobank and applying it to All of Us. Here the key issue is representativeness: the data used to estimate residual distributions in PredInterval, and the calibration set used by CalPred, must correspond to the test distribution on which coverage is evaluated. If these subsets come from a different distribution than the evaluation set, interval calibration can fail.

Accordingly, under potential distribution shift, preprocessing must ensure that the held-out subset used by PredInterval for residual computation and the calibration set used by CalPred are drawn from, or otherwise closely match, the target test distribution. Practically, this can be achieved by reserving an appropriate portion of the target dataset for held-out/calibration, which is kept separate from final evaluation, or by using a calibration cohort that is demonstrably comparable to the intended deployment population.

#### Broader benefits of normalization beyond contextual calibration

Proper phenotype normalization and data preprocessing is not only a straightforward way to improve contextual calibration; it also supports the validity of upstream analyses that precede interval construction. In particular, standard GWAS association models and most PGS construction methods implicitly assume homogeneous, well-behaved residual noise after covariate adjustment. When residual variance differs across contexts or nonlinear mean structure remains, association testing can lose power, and PGS effect estimates can be less stable – issues that can then propagate into prediction-interval construction. By addressing nonlinear mean effects and, when needed, heteroskedasticity during preprocessing, normalization can simultaneously improve association power, PGS accuracy, and the reliability of uncertainty quantification.

### Covariate-aware PredInterval to achieve contextual calibration in the absence of proper data normalization

For the rarer cases that proper data normalization was not conducted, we previously developed a covariate-aware version of PredInterval that incorporates covariates by requiring, as input, a covariate file used during model fitting. Detailed fitting instructions are provided in the software manual on GitHub, with additional technical details in the code annotations and on GitHub. Here we describe the method more formally and explain how PredInterval can be applied in settings where standard phenotype normalization has not been performed.

We use the same notations as in Xu et al.. Let *Y*_*i*_ denote the phenotype, PGS_*i*_ the polygenic score, and *z*_*i*_ ∈ ℝ^*M*^ a *M*-vector of covariates for individual *i*. The training set of size *N* is partitioned into *K* equal-sized disjoint folds *S*_1_,…,*S*_*K*_. For each individual *i*, let *k*(*i*) denote the fold to which individual *i* belongs, and let 𝒯_*k*_ = -*S*_*k*_ = {1,…,,*n*} \ *S*_*k*_ denote the (*K*-1) training folds used when fold *k* is held out.

We rely on a two-stage heteroscedastic linear regression, also known as the location-scale linear regression, to account for both mean and variance effects from covariates. Specifically, for each fold *k* = 1,…, *K*, the following two stages are fitted on 𝒯_*k*_:

Stage 1: Mean model

We fit a linear regression of the phenotype on the PGS and covariates:

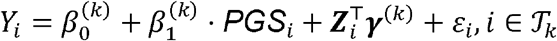

where 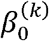 is the intercept; 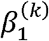 is the coefficient representing the effect of the PGS on the phenotype; ***γ*** ^(*k*)^is the corresponding *M*-vector of covariate coefficients; and *ε*_*i*_ is the residual error term, assumed to have mean zero. The fitted values, representing the model-predicted conditional mean of the phenotype for individual *i*, are given by: 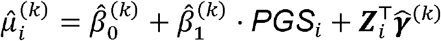. The corresponding absolute residuals, which capture the magnitude of deviation between the observed and predicted phenotype values, are then computed as: 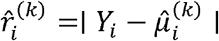 for These absolute residuals serve as the response variable in Stage 2, where they are used to model individual-level variability in prediction error as a function of the PGS and covariates.

Stage 2: Variance model

We fit a Gamma generalized linear model (GLM) with log link, we regress the absolute residuals from Stage 1 on the PGS and covariates:

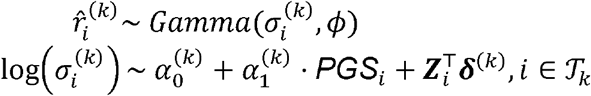

where 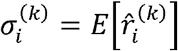 is the conditional mean of the absolute residual for individual *i*, serving as an estimate of the individual-level standard deviation; *ϕ* is the dispersion parameter of the Gamma distribution, assumed constant across individuals; 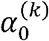 is the intercept; 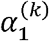is the coefficient capturing the association between the PGS and residual dispersion; and ***δ***^k^ is the corresponding *M*-vector of covariate coefficients. The log link ensures that the fitted values 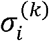 remain strictly positive, making them valid estimates of scale. These fitted values are subsequently used to standardize the nonconformity scores in the conformal prediction framework. With the above model, we obtain fitted standard deviation estimates 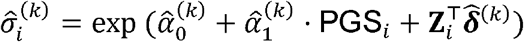. Note that we use the absolute values of the residuals rather than their squared values, and fit the standard deviation rather than the residual variance, to facilitate the computation of the standardized nonconformity score in the following steps, which takes the form of a normalized absolute residual.

Afterwards, for each held-out individual *i* ∈ *S*_*k*_, we apply the fold-*k* models to compute the predicted mean and standard deviation:

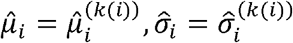

The standardized nonconformity score is then computed as:

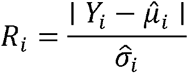

Because 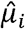 and 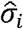 are computed using models that never saw individual *i* during fitting (since *i* ∈ *S*_*k*_ and models are fitted on 𝒯 _*k*_), these scores are honest in the leave-fold-out sense. The standardization by 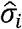 renders the scores approximately homoscedastic across individuals with different covariate profiles, enabling valid pooling across all *K* folds.

For a test individual *N* + 1 with PGS and covariates (PGS _*n*+ 1_ z_*n*+1_), we can predict the mean and standard deviation by averaging across the *K* fold-specific models:

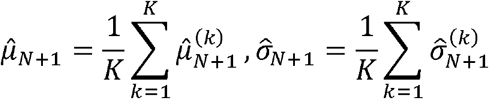

where 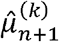 and 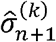 are the predictions from the fold-*k* mean and variance models respectively, evaluated at the fold-*k* PGS score for the test individual.

The (1 − *α*) prediction interval is then:

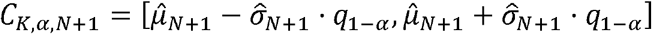

where *q*_1-*α*_ is the (1 − *α*) empirical quantile of the pooled standardized nonconformity scores 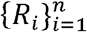:

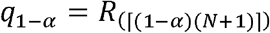

with *R* _(1)_ ≤ *R*_(2)_ ≤ ⃛ ≤ *R*_(*N*)_ denoting the order statistics of {*R*_*i*_}.

This construction inherits the marginal coverage guarantee of PredInterval, namely that *P* (*Y*_*N+*1_ ∈ *C*_*K, α,N*+1_) ≥ 1 − *α* while adaptively widening or narrowing intervals for individuals with high or low predicted variance 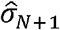.

### Contextual calibration with PredInterval when training and test populations differ

In Case III described above, when the training and test data are drawn from different populations – for example, when a PGS model is trained on one cohort and applied to another – the distribution of phenotypes, PGS values, and covariates may differ between the two samples. In this setting, the nonconformity scores 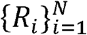 computed from the training set may no longer be exchangeable with the test nonconformity score *R*_*N*+1_, which is the key assumption underlying the coverage guarantee of PredInterval. As a result, contextual calibration may fail even when covariates are incorporated into the fitting procedure as described above.

In this setting, it is important to reserve a subset of the test data with known phenotypes as a held-out set ℋ, from which the nonconformity scores are computed. Since ℋ is drawn from the same population as the test set, the resulting nonconformity scores are exchangeable with the test scores, thus restoring the coverage guarantee. We previously extended PredInterval to automate this workflow (see the GitHub software manual for detailed instructions): the user just needs to provide a phenotype file for a subset of test individuals, and the software automatically identifies these individuals as ℋ. In practice, we found the size of the held-out set |ℋ| = 5,000 is sufficient for UK Biobank applications.

With this setup, the two-stage heteroscedastic model is then applied to ℋ for each fold *k*, using the fold-*k* PGS. Unlike the training-based procedure where the model is fitted on 𝒯_*k*_ and evaluated on the held-out fold 𝒮_*k*_, here both stages are fitted and evaluated on the same set ℋ. The nonconformity scores are then pooled across all *K* folds, giving *K* × |ℋ| scores in total. Pooling across folds increases the effective holdout sample size and stabilizes the empirical quantile estimate, at no additional computational cost since the *K* model fits are already required to predict for the remaining test individuals. The predicted mean 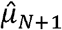 and standard deviation 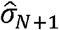 for remaining test individuals are averaged across the *K* fold-specific models as before, and the prediction interval is constructed in the same form as above with *q*_1-*α*_ now taken as the ⌈ (1 − α (*K*|ℋ|+1) ⌉order statistic of the pooled nonconformity scores. Individuals in ℋ are excluded from the prediction output since their phenotypes are observed; in the software output these individuals receive ‘NA’ for all interval columns.

### Assess contextual calibration in UK Biobank

We applied two strategies, including (i) data normalization alone and (ii) incorporating covariates directly into PredInterval, to illustrate the context-specific coverage of PredInterval using real data from the UK Biobank. We followed the same data processing strategy as in Xu et al., using 80% of the data for training and 20% for testing.

Under the data normalization approach, we directly used our original results as the covariate effects had already been removed in our original real-data analysis reported in Xu et al. through data normalization. Specifically, phenotypes were regressed on covariates (sex, age, age^2^, genotyping array, and the top 20 principal components), and the residuals were then quantile normalized. This procedure effectively implements the first strategy described above. We therefore used the previous PredInterval fitting results in Xu et al. directly and refer to this approach as *PredInterval with PGS only*.

Under the second approach, we incorporated covariates directly into PredInterval and added an additional step to train the covariate model within the training portion of the training data, as described in the previous section. We refer to this approach as *PredInterval with PGS + Covariates*.

For all methods, we computed the mean prediction coverage rate in the test set after stratifying individuals by sex (two strata) or age (three strata of approximately equal size).

The real-data results show that proper data normalization (i.e. *PredInterval with PGS only*) is sufficient to achieve contextual calibration for most analyzed traits in UK Biobank. For the trait FVC, which exhibits minor miscalibration between sexes, we conducted additional inverse normalization within each sex as explained in Case II in a previous section, which leads to contextual calibration without the use of covariates.

Finally, to illustrate the consequences of inadequate covariate adjustment, we considered scenarios in which data normalization is not properly conducted. Specifically, we generated reconstructed phenotypes by adding back the original standardized effects of sex, age, and age^2^ to the normalized phenotypes. This allowed us to systematically manipulate the covariate effects and examine their impact on contextual calibration. We considered two scenarios: (i) Same Covariate Effect, where covariate effects are added back consistently across all samples, representing a situation where covariate effects are present but uniform across training and test samples and not properly accounted for in the model; and (ii) Shifted Covariate Effect, where covariate effects are added back with a larger magnitude, which is twice as large in the test set as in the training set, thus creating a deliberate mismatch between the training and test distributions. These scenarios mimic real-world situations in which covariate effects may be incompletely controlled or differ across populations or study designs in the absence of proper data normalization.

We evaluated both PredInterval with PGS Only and PredInterval with PGS + Covariates in each scenario. For the Same Covariate Effect setting, PredInterval with PGS + Covariates was applied in the standard way, fitting the two-stage heteroscedastic model using the training CV folds as described above. For the Shifted Covariate Effect setting, where the covariate distribution differs between training and test populations, the standard training-based nonconformity scores are no longer exchangeable with the test scores. We therefore applied PredInterval with PGS + Covariates using a held-out subset of the test data with known phenotypes as the holdout set ℋ, as described in the previous section. By comparing results across these settings, we demonstrate how improper normalization or unmodeled covariate effects can lead to context-specific miscalibration, and how proper data normalization or explicit incorporation of covariates, with appropriate holdout set selection, can restore contextual calibration under both uniform and shifted covariate effects.

### Contextual calibration in the simulations of Hou et al. and Shi et al

Because we did not observe contextual miscalibration in real-data analyses, we carefully examined why Hou et al. and Shi et al. observed contextual miscalibration for other methods in their simulations. Both studies simulate phenotypes under extreme contextual heterogeneity using a data-generating mechanism that is fully consistent with the parametric CalPred model. We focus on the most extreme setting in Shi et al., which uses 60,000 individuals and 82,204 publicly available SNPs. We followed their procedure and used their code to simulate two settings: *h*^2^ = 0.5 and *h*^2^ = 0.05.

In each replicate, we first simulated a contextual variable for each individual from a standard normal distribution. We then simulated heteroskedastic residual for each individual from a normal distribution with individual-specific variance modeled as an exponential function of the context variable. Next, we simulated SNP effect sizes for all 82k SNPs from a normal distribution and multiplied them by genotypes to form the genetic value, which was then rescaled to have mean 0 and standard deviation 1. The phenotype was generated by combining the genetic value and heteroskedastic residuals: for the *h*^2^ = 0.5 setting, the phenotype was formed by summing these two together with equal weights; for the *h*^2^ = 0.05 setting, the components were reweighted so that genetic effects explained 5% of the phenotypic variance. Finally, the 60,000 individuals were split into a development sample of 50,000, with 40,000 as training and 10,000 as held-out, and a test sample of 10,000.

For each replicate, we first analyzed the unnormalized phenotype using the original GWAS and polygenic-score pipeline with PRS-CS. We then normalized the phenotype using a model fit on the training set, reran GWAS and PGS construction, and compared marginal association statistics, PGS accuracy, and prediction-interval performance in the test set before versus after normalization. In the *h*^2^ = 0.05 setting, we applied the two-stage regression procedure described in Case II above for normalization. In the *h*^2^ = 0.5 setting, because genetic effects are unrealistically large, are homoskedastic and independent of context, and heteroskedasticity is introduced only through the residual component, we modified the first-stage mean adjustment to include the PGS constructed from the unnormalized phenotype. This yields residuals that are approximately free of genetic signal for the variance-modeling step. We then fit the second-stage regression of log (*ê*^2^)on the contextual variable to estimate the heteroskedastic scale, and finally reconstructed the normalized phenotype by combining the variance-stabilized residual component with the PGS contribution.

For evaluation, we applied both *PredInterval with PGS only* and *PredInterval with PGS + Covariates* in the test sample to assess (i) association signals as summarized by the median *χ*^2^ statistic across causal SNPs, (ii) PGS accuracy, as measured by correlation and root of mean squared error (RMSE) between PGS and the true genetic value, and (iii) prediction-interval calibration, comparing results before and after normalization.

## Data Availability

Individual-level UK Biobank data used in this study cannot be shared publicly by the authors due to participant privacy and data-use restrictions. These data are available to approved researchers through the UK Biobank access process under standard terms. This study was conducted using the UK Biobank resource under Application Number 30686. Additional data generated or analyzed in this study are contained in the manuscript and supplementary materials.

## Acknowledgements

This study was supported by the National Institutes of Health (NIH) grants R01HG009124. The funders had no role in study design, data collection and analysis, decision to publish or preparation of the manuscript. This study has been conducted using UK Biobank resource under Application Number 30686. UK Biobank was established by the Wellcome Trust medical charity, Medical Research Council, Department of Health, Scottish Government and the Northwest Regional Development Agency. It has also had funding from the Welsh Assembly Government, British Heart Foundation and Diabetes UK.

## Code availability

PredInterval, along with analysis code to reproduce the results presented in the present study, is available at https://github.com/xuchang0201/PredInterval.

## Contributions

X.Z. conceived the study and developed the methods. X.Z. and S.H. implemented the software. C.X. and S.H. conducted the experiments and analyzed and interpreted the data. X.Z. drafted the manuscript, with revisions from C.X. and S.H. All authors critically reviewed the manuscript, provided feedback, and approved the final version.

