## Supplementary Note for "Calibrated Prediction Intervals for Polygenic Scores: Updated Comparisons, Contextual Calibration, and Data Normalization"

**1. Investigation of our previous use of CalPred in Xu et al.**

Our previous use of CalPred, as reported in Xu et al., followed directly from the CalPred software manual, with supporting evidence from the "Simulation studies of context-specific calibration" and "Real data analysis" subsections of the CalPred Methods section. Given that methods can be implemented differently across software packages, following the manual is the most natural course of action for users. The manual described software usage with two data parts, and these subsections specified detailed sample sizes for those two parts accordingly; we therefore adopted the provided example code from GitHub directly and reasonably assumed that any additional data splitting was handled internally by the software, especially given that the calibration set size is recommended to be only 500 in the “Constructing calibrated and context-specific prediction intervals” subsection of the CalPred Methods section.

This assumption was further reinforced by three observations: first, we were able to run the example data and reproduce the main simulation results reported in the CalPred paper, which employs a notably different simulation strategy than the more realistic simulations in Xu et al.; second, we formulated and validated a hypothesis, confirmed through additional simulations and detailed below, that reconciled the differences observed between our simulations and those reported in the CalPred paper; and third, our real data results were consistent with those reported in the CalPred paper, and real data performance is arguably the more meaningful benchmark. Together, these three observations gave us confidence that we were applying the software correctly.

Our working hypothesis at that time was that CalPred's algorithm becomes more accurate with larger sample sizes due to improved estimation of SNP effect sizes, such that small sample sizes would produce downward bias in coverage. This hypothesis coherently explained two sets of discrepancies. First, it accounted for the difference between our simulation and real data results, as real data involved larger sample sizes. Second, it explained the difference between our simulations and those reported in the CalPred paper. Specifically, the CalPred simulations generate PGS by drawing directly from a normal distribution, rather than simulating SNP effect sizes, multiplying onto real genotype data, and estimating SNP effects to construct the PGS as we did, which is a strategy effectively equivalent to assuming knowledge of the true SNP effect sizes. Indeed, when we reran our simulations supplying the true SNP effect sizes directly, we also observed unbiased results, consistent with this hypothesis. In hindsight, this hypothesis was incorrect in light of the new evidence regarding the three-way data split; however, because it coherently explained all discrepancies observed at the time, it gave us reasonable confidence that our software usage was correct.

We subsequently learned from the team of Shi et al. that the intended use of CalPred requires users to manually split the input data into three sets, which is a detail not made explicit in the software manual. We have since updated our CalPred results accordingly. As reported in the manuscript, corrected usage substantially improves simulation performance but does not substantially improve real data performance on its own, which remains downward biased. Further investigation reveals that a calibration set approximately ten times larger than the 500-sample default recommended in the "Constructing calibrated and context-specific prediction intervals" subsection of the CalPred Methods section is needed to achieve coverage centered on the targeted level in real data. Corrected usage is therefore necessary but not sufficient for strong real data performance; adequate calibration set size is an additional and independent requirement.

The main conclusions and contributions of PredInterval remain intact. It continues to offer a non-parametric approach to achieving calibrated prediction intervals on the original PGS, that, by virtue of its use of prediction residuals, can be easily extended in a variety of directions. The updated results do, however, show that CalPred is an equally effective parametric alternative in terms of prediction coverage when a sufficiently large calibration dataset is used.
